# Neuropsychiatric Symptoms Cluster and Fluctuate Over Time in Behavioral Variant Frontotemporal Dementia

**DOI:** 10.1101/2024.09.26.24314180

**Authors:** Christopher B. Morrow, Vidyulata Kamath, Bradford C. Dickerson, Mark Eldaief, Neguine Rezaii, Bonnie Wong, Scott McGinnis, Ryan Darby, Adam M. Staffaroni, Maria I. Lapid, Belen Pascual, Julio C. Rojas, Joseph C. Masdeu, Kyrana Tsapkini, Edward D. Huey, Daniel W. Fisher, Alexander Pantelyat, Akshata Balaji, Eric Sah, Irene Litvan, Katya Rascovsky, Nupur Ghoshal, Kimiko Domoto-Reilly, John Kornak, Chiadi U. Onyike, the ALLFTD Consortium

## Abstract

**Objectives:** Cognitive and behavioral phenomena define behavioral variant frontotemporal dementia (bvFTD), but neuropsychiatric symptoms (NPS) outside the core criteria are common throughout the illness. Identifying how NPS cluster in bvFTD may clarify the underlying neurobiology of bvFTD-related NPS and guide development of therapies.

**Methodology:** Participants (N=354) with sporadic and genetic bvFTD were enrolled in the ARTFL LEFFTDS Longitudinal Frontotemporal Lobar Degeneration Consortium. Dementia stage was defined as early (CDR® plus NACC FTLD ≤ 1) or advanced (CDR® plus NACC FTLD ≥ 1). Baseline and annual follow-up visit data were analyzed to compare NPS across stages of bvFTD. Psychiatric states were captured using the Neuropsychiatric Inventory-Questionnaire and Clinician Judgement of Symptoms. Polychoric cluster analysis was used to describe NPS clusters.

**Results:** NPS were highly prevalent (≥ 90%) in early and late bvFTD. Four NPS clusters were identified based on magnitude of factor loadings: affective, disinhibited, compulsive, and psychosis. Neuropsychiatric symptoms fluctuated across visits. In the affective cluster, depression and anxiety showed the least visit-to-visit stability. In the disinhibited cluster, elation showed the least stability. Symptoms in the psychosis and compulsive clusters (hallucinations, delusions, obsessions/compulsions, and hyperorality) were largely stable, persisting from visit-to-visit in more than 50% of cases.

**Conclusion:** NPS in bvFTD are frequent and cluster into four discrete groups in bvFTD. These clusters may result from specific neural network disruptions that could serve as targets for future interventions. The fluctuating nature of NPS in bvFTD suggests that they are not reliable markers of disease progression or stage.

## Introduction

The behavioral variant of frontotemporal dementia (bvFTD) is a clinically and pathologically heterogeneous syndrome defined by specific cognitive and behavioral phenomena.(1) However, neuropsychiatric symptoms (NPS) outside the core diagnostic criteria, like depression, elation, hallucinations, and delusions, are common at every stage of the disease. The common bvFTD-related NPS can overlap with those seen in primary psychiatric disorders (PPD), often obscuring diagnostic clarity.(2–4). Understanding how NPS emerge and cluster in bvFTD could improve the accuracy of early diagnosis and inform treatment strategies.

There is preliminary evidence of psychiatric symptom clusters in bvFTD, but the validity and stability of these clusters across diverse patient cohorts has not been established.(5, 6) Identifying reliable NPS clusters in bvFTD may provide insights into the neurobiology of NPS in bvFTD as well as in related neurodegenerative disorders like Alzheimer disease (AD) and dementia with Lewy bodies (DLB), where psychiatric symptoms are similarly prevalent.(7) Better understanding of NPS clusters in bvFTD could also inform treatment strategies, improving quality of life and function for patients living at all stages of bvFTD. Furthermore, understanding how psychiatric symptoms relate to disease stage may provide useful information regarding the utility of NPS for bvFTD staging and monitoring. Identifying early symptoms and biomarkers of bvFTD is essential for optimizing recruitment for clinical trials of emerging disease-modifying therapies.(8, 9) If NPS correlate with bvFTD stage, they will provide helpful staging information; however, if they fluctuate in bvFTD as they do in PPD or in AD, they are not likely to be reliable markers of disease progression.(10)

Given this background, our primary aim was to identify NPS clusters in bvFTD based on symptom correlations. Using cluster analysis techniques, we tested the hypothesis that NPS clusters in bvFTD overlap with constructs commonly observed in PPD. A complementary aim of this study was to evaluate the stability of specific NPS over time. We tested the hypothesis that specific NPS fluctuate at different rates over time in bvFTD and may not reliably associate with neurodegeneration.

## Methods

### Participants

Participants were enrolled in the ARTFL LEFFTDS Longitudinal Frontotemporal Lobar Degeneration (ALLFTD) study. Participants underwent extensive clinical assessment. The study protocol and procedures can be found in earlier papers.(11, 12) Participants with a primary clinical diagnosis of bvFTD who met formal criteria for probable bvFTD at one or more study visits were included in the analyses; those not meeting criteria for probable bvFTD or having a primary diagnosis other than bvFTD were excluded.(1) Pathological confirmation of bvFTD diagnosis was not available. Disease severity was defined based on Clinical Dementia Rating (CDR®) plus NACC FTLD Behavior & Language Domains global score (CDR® plus NACC FTLD).(13, 14) Participants with CDR® plus NACC FTLD scores of ≤1 at visit 1 were classified as early-stage, and those with CDR® plus NACC FTLD scores of 2 or 3 as advanced-stage.

### Clinical Assessment

Data from the Neuropsychiatric Inventory Questionnaire (NPI-Q) captured the following neuropsychiatric symptoms: depression, anxiety, hallucinations, delusions, agitation, apathy, disinhibition, irritability, and elation. We utilized these NPS as they correspond most closely with symptoms in common PPD including major depressive disorder, bipolar disorder, and schizophrenia. The NPI-Q is a validated and widely used informant-rated scale to assess NPS in dementia syndromes, including bvFTD.(15–19) The aforementioned NPS were analyzed as dichotomous variables based on their presence or absence on the NPI-Q. The hyperorality and ritualistic/compulsive behavior variables were drawn from Uniform Data Set (UDS) version 3 Form B9F – Clinical PPA and bvFTD Features, part of the FTLD Module.(20) This module was implemented by the NIA Alzheimer’s Disease Research Centers (ADRC) to help differentiate the neuropsychological characteristics of FTD from AD. The FTLD module includes a series of psychometric tests sensitive to the behavioral and language impairments common in FTD syndromes (i.e., bvFTD and primary progressive aphasia) and is described in detail in earlier studies.(21) Hyperorality and ritualistic/compulsive behavior variables were recorded as present if marked as “definitely present” and absent otherwise. Cognitive and functional ability were assessed using the Montreal Cognitive Assessment (MoCA) and the CDR® plus NACC FTLD sum of boxes score.(22, 23) We examined caregiver burden using the Zarit Burden Interview, a 22-item instrument that is well validated for capturing caregiver burden in FTD spectrum disorders.(24) Functional ability was assessed using the Functional Activities Questionnaire (FAQ).

### Statistical Methods

Differences in participant characteristics and clinical outcomes were compared using two-sided t-tests for continuous variables and Pearson χ2 tests for categorical variables. A sensitivity analysis including only those NPI-Q scores that were moderate or severe was conducted. A sensitivity analysis including cases of “questionable” hyperorality and ritualistic/compulsive behavior was also conducted.

We performed an exploratory factor analysis of NPS to determine whether specific symptoms could be psychometrically grouped into distinct factors. To determine the appropriate number of factors, a polychoric correlation matrix of all eleven NPS (hallucinations, delusions, agitation, depression, anxiety, elation, apathy, disinhibition, irritability, hyperorality, and ritualistic/compulsive behavior) was evaluated in a parallel analysis for principal components. To evaluate the robustness of the principal components analysis (PCA), we generated 1000 bootstrap samples from the original dataset and performed a PCA on each sample to examine for variability in the principal components. Polychoric correlations, rather than Pearson correlations, were used because the NPS measures included missing values and were not normally distributed (i.e. non-parametric).(25) Parallel analysis consists of randomly generating a number of simulated data sets (conventionally 1000) with dimensions, means, and standard deviations identical to those in the observed data but without intrinsic relationships between variables as the data are randomly generated.(26) The simulated datasets undergo PCA, and the means of each eigenvalue are calculated. The ideal number of factors is chosen such that eigenvalues in the observed data are greater than the respective mean eigenvalues from the simulated data sets. (27) We inspected a scree plot and confirmed that the number of factors identified using the parallel analysis matched those identified on the scree plot. A scree plot consists of a graphical representation of eigenvalues with the appropriate number of factors typically occurring at a point where the decrease in eigenvalue with additional factors levels off. If the parallel analysis was ambiguous (i.e., the observed eigenvalue was only slightly higher (<0.05) than the simulated eigenvalue) and the scree plot favored a smaller number of factors, we selected the smaller number of factors.

Once the appropriate number of factors was selected, a factor analysis was performed. Factor loadings were rotated using the promax rotation, allowing for correlations among the factors. Specific NPS were excluded if they had high unique variance (uniqueness > 0.6) as high unique variance indicates that a significant amount of the variance is complementary to that of the other variables. This improves the model’s overall fit and factor structure. (28)

A final model was fit using the optimal number of factors with the final set of NPS. The factor loadings were rotated using the promax rotation. These analyses were performed for the entire participant population across all visits, as well as cross-sectionally using baseline visit data in early-stage and advanced-stage participant groups, respectively. Individual NPS with loadings close to +1.00 or –1.00 were interpreted as loading strongly onto a factor, while those nearest zero were considered as loading weakly onto a factor. Given the exploratory nature of factor analysis, no statistical threshold was set to determine the adequacy of factor loading. However, loadings of 0.3 or higher are generally considered to be salient, and we interpreted loadings of 0.5 or higher as constituting a meaningful (moderately high) association between a variable and a factor.(29).

The stability of NPS overtime was assessed by calculating the proportion of NPS that either resolved or persisted from one study visit to the subsequent visit. Visits occurred at approximately annual intervals although there was variability across participants (mean of 463 days between visits). Disease progression was considered present if a participant’s CDR® plus NACC FTLD increase by 1 or more points over the course of the study.

The statistical significance level, a, was set at 0.05. STATA SE 17 (StataCorp LP, College Station, TX) was used for all analyses.

## Results

### Demographics

Demographic data are shown in Table 1. Of 1,316 participants with baseline data, 354 participants had a primary clinical diagnosis of bvFTD. Of the 354 participants with a baseline visit, there were 109 participants with one follow-up visit, 45 participants with two, 24 participants with three, 12 participants with four, and 5 participants with five follow-up visits. The mean number of visits in the study was 2.2 with a standard deviation of 1.5 visits. There were 145 participants classified as early-stage (22 with a CDR® plus NACC FTLD of 0.5) and 209 classified as advanced stage at the baseline visit. As expected, mean MoCA scores were lower in the advanced-stage participants (15.3 versus 21.6, p<0.001) and CDR® FTLD-SoB scores were higher (12.1 versus 44.9, p<0.001). There was a higher proportion of *C9orf72* mutation carriers in the advanced-stage group than the early-stage group (18.2% versus 7.6%, p = 0.01). There were no other differences in age, sex, education, or gene status between the early-stage and advanced-stage groups.

**Table 1:** Baseline Demographic Characteristics and Mutation Status for bvFTD Patients.

|  | All Participants<br>n = 354 | Early Stage<br>(CDR® FTLD ≤ 1)<br>n = 145 | Advanced Stage<br>(CDR® FTLD > 1)<br>n = 209 | p-value |
| --- | --- | --- | --- | --- |
|  |  | mean (SD) | mean (SD) |  |
| Age (years) | 61.9 (9.2) | 61.9 (7.8) | 61.8 (10.0) | 0.95 |
| Sex (% Men) | 56.2 | 60.7 | 53.1 | 0.16 |
| Education level (years) | 15.8 (2.6) | 16.0 (2.6) | 15.6 (2.6) | 0.11 |
| MoCA | 18.0 (7.0) | 21.6 (5.2) | 15.3 (7.0) | <0.001 |
| CDR® FTLD -SOB Score (mean) | 9.2 (525.1) | 4.9 (292.0) | 1212.1 (44.4) | <0.001 |
| *C9orf72* (n, %) | 49 (13.8) | 11 (7.6) | 38 (18.2) | 0.01 |
| *GRN* (n, %) | 23 (6.5) | 13 (9.0) | 10 (4.8) | 0.05 |
| *MAPT* (n, %) | 31 (8.8) | 16 (11.0) | 15 (7.2) | 0.07 |
| Sporadic (n, %) | 174 (49.2) | 67 (46.2) | 107 (51.2) | 0.24 |

\*t-values for continuous variables, $\chi^2$ for binary variables
\*\*genetic status unknown in 21.8% of participants

### Baseline Neuropsychiatric symptoms

The frequency of NPS at baseline is displayed in Table 2. Neuropsychiatric symptoms were common in early and advanced-stage participants. Apathy, irritability, and disinhibition were the most common NPS at baseline—occurring in 74%, 63%, and 64% of participants, respectively. Within the early-stage group, apathy was less common in MAPT mutation carriers than in GRN mutation carriers (38% [95% CI 14%-61%] versus 77% [95% CI 54%-99%], p-value 0.03) and sporadic bvFTD (38% [95% CI 14%-61%] versus 83% [95% CI 74%-92%], p-value <0.001). Obsessions/compulsions were also less common in early-stage MAPT mutation carriers than in sporadic bvFTD (8% [95% CI 0%-24%] versus 52% [95% CI 40%-64%], p-value 0.005). Within the advanced-stage group, disinhibition was less common among MAPT mutation carriers compared to C9orf72 mutation carriers (33% [95% CI 7%-60%] versus 81% [95% CI 68%-93%], p-value 0.002) and sporadic bvFTD (33% [95% CI 7%-60%] versus 75% [95% CI 67%-83%], p-value 0.003). Hyperorality was more common in advanced stage sporadic bvFTD than in C9orf72 mutation carriers (73% [95% CI 64%-81%] versus 50% [95% CI 34%-66%], p-value 0.01) and MAPT mutation carriers (73% [95% CI 64%-81%] versus 47% [95% CI 21%-72%], p-value 0.04). Hyperorality and obsessions/compulsions were more common in advanced-stage participants compared to early-stage participants (63% [95% CI 56%-70%] versus 50% [95% CI 42%-58%], p-value 0.02 and 70% [95% CI 64%-76%] versus 46% [95% CI 37%-54%], p-value <0.001) respectively. Depression was more common in early-stage participants compared to advanced-stage participants (49.3% [95% CI 41%-58%] versus 29.0% [95% CI 23%-35%], p-value <0.001). A higher proportion of participants with GRN mutations progressed (change in CDR® plus NACC FTLD ≥ 1) during the study than those with C9orf72 mutations (75% [95% CI 54%-96% versus 29% [95% CI 13%-45%], p-value 0.003), and sporadic bvFTD (75% [95% CI 54%-96% versus 36% [95% CI 18%-53%], p-value 0.01). At the baseline visit, the majority (>70%) of participants experienced 3 or more NPS concurrently (Table 3).

**Table 2:** Percent frequency of individual neuropsychiatric symptoms at baseline visit.

| NPS | Early Stage<br>(CDR® FTLD ≤ 1) |  |  |  |  | p-value* | Advanced Stage<br>(CDR® FTLD > 1) |  |  |  |  | p-value* |
| --- | --- | --- | --- | --- | --- | --- | --- | --- | --- | --- | --- | --- |
|  | All<br>(n = 145) | *Sporadic*<br>(n = 67) | *GRN*<br>(n = 13) | *MAPT*<br>(n = 16) | *C9*<br>(n = 11) |  | All<br>(n = 209) | *Sporadic*<br>(n = 107) | *GRN*<br>(n = 10) | *MAPT*<br>(n = 15) | *C9*<br>(n = 38) |  |
| Hallucinations | 5.8 | 1.9 | 7.7 | 7.7 | 11.1 | 0.49 | 7.9 | 3.3 | 11.1 | 15.4 | 9.1 | 0.26 |
| Delusions | 18.1 | 17.2 | 7.7 | 25.0 | 20.0 | 0.67 | 18.4 | 16.4 | 20.0 | 15.4 | 19.0 | 0.98 |
| Agitation | 48.2 | 58.5 | 38.5 | 37.5 | 20.0 | 0.07 | 54.0 | 58.7 | 50.0 | 53.9 | 54.1 | 0.92 |
| Depression | 49.3 | 53.1 | 53.9 | 31.3 | 40.0 | 0.41 | 29.0 | 31.1 | 30.0 | 0.0 | 27.8 | 0.13 |
| Anxiety | 48.9 | 48.4 | 38.5 | 37.5 | 40.0 | 0.80 | 47.5 | 49.0 | 70.0 | 33.3 | 40.5 | 0.28 |
| Apathy | 73.6 | 82.8 | 76.9 | 37.5 | 70.0 | **0.003** | 79.7 | 79.1 | 90.0 | 50.0 | 78.4 | 0.10 |
| Elation | 27.9 | 28.2 | 38.5 | 37.5 | 20.0 | 0.75 | 24.4 | 22.9 | 30.0 | 0.0 | 33.3 | 0.12 |
| Disinhibition | 64.3 | 71.9 | 61.5 | 62.5 | 60.0 | 0.49 | 71.9 | 75.0 | 50.0 | 33.3 | 80.6 | **0.005** |
| Irritability | 62.6 | 70.3 | 46.2 | 62.5 | 40.0 | 0.15 | 56.2 | 58.1 | 60.0 | 38.5 | 62.2 | 0.51 |
| Hyperorality | 50.0 | 56.7 | 30.0 | 25.0 | 33.3 | 0.08 | 63.0 | 72.6 | 70.0 | 46.7 | 50.0 | **0.03** |
| Obsessions<br>/Compulsions | 45.6 | 52.2 | 20.0 | 8.3 | 22.2 | **0.008** | 70.2 | 75.5 | 40.0 | 73.3 | 73.7 | 0.12 |

\* chi-square comparison by gene category within disease stage

**Table 3:** Percent frequency of individual NPS at baseline visit by CDR® FTLD score.

| NPS | CDR® FTLD |  |  |  |  | p-value* |
| --- | --- | --- | --- | --- | --- | --- |
|  | 0<br>(n = 6) | 0.5<br>(n = 22) | 1<br>(n = 117) | 2<br>(n = 176) | 3<br>(n = 33) |  |
| Hallucinations | 20.0 | 4.5 | 5.3 | 8.0 | 7.4 | 0.72 |
| Delusions | 33.3 | 9.1 | 19.1 | 18.7 | 16.7 | 0.69 |
| Agitation | 16.7 | 36.4 | 52.2 | 54.7 | 50.0 | 0.23 |
| Depression | 33.3 | 45.5 | 50.9 | 30.6 | 20.0 | **0.002** |
| Anxiety | 33.3 | 36.4 | 52.3 | 43.0 | 75.0 | **0.02** |
| Apathy | 16.7 | 68.2 | 77.7 | 79.2 | 82.8 | **0.006** |
| Elation | 16.7 | 22.7 | 29.5 | 24.3 | 25.0 | 0.85 |
| Disinhibition | 16.7 | 54.6 | 68.8 | 75.3 | 51.7 | **0.002** |
| Irritability | 33.3 | 59.1 | 64.9 | 57.2 | 50.0 | 0.35 |
| Hyperorality | --- | 31.6 | 53.0 | 59.4 | 81.8 | **0.002** |
| Obsessions /Compulsions | --- | 5.3 | 52.1 | 68.6 | 78.8 | **<0.001** |
| **Total # of NPS** |  |  |  |  |  |  |
| 0 | 50.0 | 0.0 | 1.7 | 3.4 | 0.0 |  |
| 1-2 | 16.7 | 27.3 | 15.4 | 12.5 | 12.1 | **<0.001** |
| 3+ | 33.3 | 72.7 | 82.9 | 84.1 | 87.9 |  |

\*t-values for continuous variables, $\chi^2$ for binary variables

### Exploratory Factor Analysis

The results of the exploratory factor analysis are displayed in Tables 4-6. The parallel analysis of the 11 NPS items supported a model with 4 factors for early-stage, advanced-stage, and all participants combined.

**Table 4:** Four-Factor NPS Cluster Model in Early-Stage FTD.

| NPS | Factor 1<br>(Affective) | Factor 2<br>(Disinhibited) | Factor 3<br>(Compulsive) | Factor 4<br>(Psychosis) |
| --- | --- | --- | --- | --- |
| Agitation | **0.84** | 0.38 | 0.22 | -0.02 |
| Depression | **0.73** | -0.02 | -0.20 | 0.04 |
| Anxiety | **0.67** | 0.21 | -0.00 | 0.01 |
| Irritability | **0.74** | 0.56 | -0.04 | 0.26 |
| Elation | 0.15 | **0.92** | 0.16 | 0.26 |
| Disinhibition | 0.61 | **0.89** | 0.19 | 0.28 |
| Hyperorality | 0.06 | 0.22 | **0.76** | 0.20 |
| Obsessions/Compulsions | -0.04 | 0.10 | **0.82** | 0.04 |
| Hallucinations | 0.06 | 0.30 | 0.13 | **0.99** |
| Delusions | 0.06 | 0.30 | 0.13 | **0.99** |

\*Factor loadings are based on a promax rotated solution. Boldface type indicates the primary (i.e., highest) factor loading for each item. NPS loadings close to +1.00 or -1.00 are interpreted as loading strongly onto a factor, while those nearest zero are considered as loading weakly onto a factor.

For the early-stage group (Table 4), the observed and simulated eigenvalues for the fourth component were 1.26 and 1.15, respectively, while the observed and simulated eigenvalues for the fifth component were 0.97 and 1.1, respectively. The scree plot supported a model with 4 or 5 factors. We selected a model with 4 factors as the parallel analysis was not ambiguous and this was the more parsimonious model (Supplementary Figure 1). After a promax rotation of the 4-factor solution, we removed apathy from the early-stage model due to high unique variance (uniqueness 0.62, loading of 0.57 in disinhibited cluster if not excluded). The final model for the early-stage included the following four factors, which we named according to the character of symptoms within each factor: Factor 1 – *Affective* (depression, anxiety, agitation, irritability); Factor 2 – *Disinhibited Type A* (elation, disinhibition); Factor 3 – *Compulsive* (obsessive/ritualistic behaviors, hyperorality); and Factor 4 – *Psychosis* (hallucinations, delusions).

For the advanced-stage group (Table 5), the observed and simulated eigenvalues for the fourth component were 1.2 and 1.1, respectively, while the observed and simulated eigenvalues for the fifth component were 0.93 and 1.1, respectively. This supported a four-factor model which was confirmed with inspection of the scree plot which also supported a model with four factors (Supplementary Figure 2). After a promax rotation of the four-factor solution, we removed elation from the advanced-stage model due to high unique variance (uniqueness 0.73, loading of 0.37 within disinhibited factor if not excluded). The final model for the advanced-stage participants included the following four factors, which we named according to the character of the symptoms within each factor: Factor 1 – *Affective* (depression, anxiety, agitation, irritability); Factor 2 – *Disinhibited Type B* (disinhibition, apathy); Factor 3 – *Compulsive* (obsessive/ritualistic behaviors, hyperorality); and Factor 4 – *Psychosis* (hallucinations, delusions).

**Table 5:** Four-Factor NPS Cluster Model in Advanced-Stage FTD.

| NPS | Factor 1<br>(Affective) | Factor 2<br>(Disinhibited) | Factor 3<br>(Compulsive) | Factor 4<br>(Psychosis) |
| --- | --- | --- | --- | --- |
| Agitation | **0.78** | 0.36 | -0.01 | 0.40 |
| Depression | **0.66** | 0.23 | -0.23 | 0.12 |
| Anxiety | **0.48** | -0.17 | 0.44 | 0.17 |
| Irritability | **0.77** | 0.17 | 0.20 | 0.29 |
| Apathy | 0.25 | **0.74** | 0.31 | 0.009 |
| Disinhibition | 0.32 | **0.70** | 0.02 | 0.19 |
| Hyperorality | -0.02 | 0.25 | **0.73** | -0.04 |
| Obsessions/Compulsions | 0.04 | 0.05 | **0.56** | -0.28 |
| Hallucinations | 0.35 | 0.10 | -0.10 | **1.0** |
| Delusions | 0.35 | 0.10 | -0.10 | **1.0** |

\*Factor loadings are based on a promax rotated solution. Boldface type indicates the primary (i.e., highest) factor loading for each item. NPS loadings close to +1.00 or -1.00 are interpreted as loading strongly onto a factor, while those nearest zero are considered as loading weakly onto a factor.

When all participants were assessed together across all visits (Table 6), the observed and simulated eigenvalues for the fourth component were 1.1 and 1.1, respectively, while the observed and simulated eigenvalues for the fifth component were 0.91 and 1.0, respectively. The scree plot supported a model with four factors, and therefore a model with four factors was selected (Supplementary Figure 3). After a promax rotation of the four-factor solution, we removed apathy due to high unique variance (uniqueness 0.68, loading of 0.57 within disinhibited factor if not excluded). The final model with all participants across all visits included the following four factors, which we named according to the character of the symptoms within each factor: Factor 1 – *Affective* (depression, anxiety, agitation, irritability); Factor 2 – *Disinhibited Type A* (disinhibition, elation); Factor 3 – *Compulsive* (obsessive/ritualistic behaviors, hyperorality); and Factor 4 – *Psychosis* (hallucinations, delusions).

The sensitivity analysis limiting NPS to only moderate or severe symptoms resulted in small changes to the derived clusters. In the model limited to baseline visit data of early-stage participants, the derived factors remained the same aside from irritability loading more strongly with the disinhibited cluster. In the model limited to baseline data of advanced-stage participants, the derived factors were identical. In the model with all participants included across all visits, a three-factor model without a disinhibited cluster best fit the data (elation, disinhibition, and apathy were excluded due to elevated uniqueness).

The sensitivity analysis including questionable cases of obsessive/ritualistic behaviors and hyperorality also resulted in small changes to the derived clusters. In the model limited to baseline visit data of early-stage participants, the derived clusters were unchanged. In the model limited to baseline data of advanced-stage participants, elation and obsessions were dropped due to elevated uniqueness, resulting in four clusters including psychosis (hallucinations, delusions), affective (depression, agitation, irritability), hyperoral (hyperorality, anxiety), and disinhibited (disinhibition, apathy). In the model with all participants included across all visits, a three-factor model best fit the data (elation, anxiety, and apathy excluded due to elevated uniqueness).

### Persistence of NPS

The persistence of specific NPS across visits is displayed in Table 7. At the first follow-up visit, NPS generally persisted in greater than 60% of cases with fewer than 30% of cases resolving. Elation was the one exception which persisted in only 55% of cases and resolved in 42% at the first follow-up. At the second follow-up visit, depression, persisted in only 39% of cases with all other NPS persisting in greater than 50% of cases. Follow-up was sparse after 3 visits, however, at the third follow-up anxiety persisted in only 36% of cases and resolved in 64%, while depression persisted in only 29% of cases and resolved in 71%. Other NPS persisted in greater than 50% of cases at the third follow-up visit.

**Table 6:** Four-Factor NPS Cluster Model Across All Visits and Disease Severity.

| NPS | Factor 1<br>(Affective) | Factor 2<br>(Disinhibited) | Factor 3<br>(Compulsive) | Factor 4<br>(Psychosis) |
| --- | --- | --- | --- | --- |
| Agitation | **0.75** | 0.41 | 0.11 | 0.18 |
| Depression | **0.62** | -0.02 | -0.14 | 0.22 |
| Anxiety | **0.62** | 0.10 | 0.20 | 0.30 |
| Irritability | **0.77** | 0.37 | 0.06 | 0.30 |
| Elation | 0.25 | **0.66** | 0.29 | 0.16 |
| Disinhibition | 0.45 | **0.65** | 0.21 | 0.15 |
| Hyperorality | 0.02 | 0.34 | **0.67** | 0.07 |
| Obsessions/Compulsions | -0.02 | 0.20 | **0.70** | -0.07 |
| Hallucinations | 0.34 | 0.12 | 0.01 | **0.99** |
| Delusions | 0.34 | 0.12 | 0.01 | **0.99** |

\*Factor loadings are based on a promax rotated solution. Boldface type indicates the primary (i.e., highest) factor loading for each item. NPS loadings close to +1.00 or -1.00 are interpreted as loading strongly onto a factor, while those nearest zero are considered as loading weakly onto a factor.

**Table 7:**
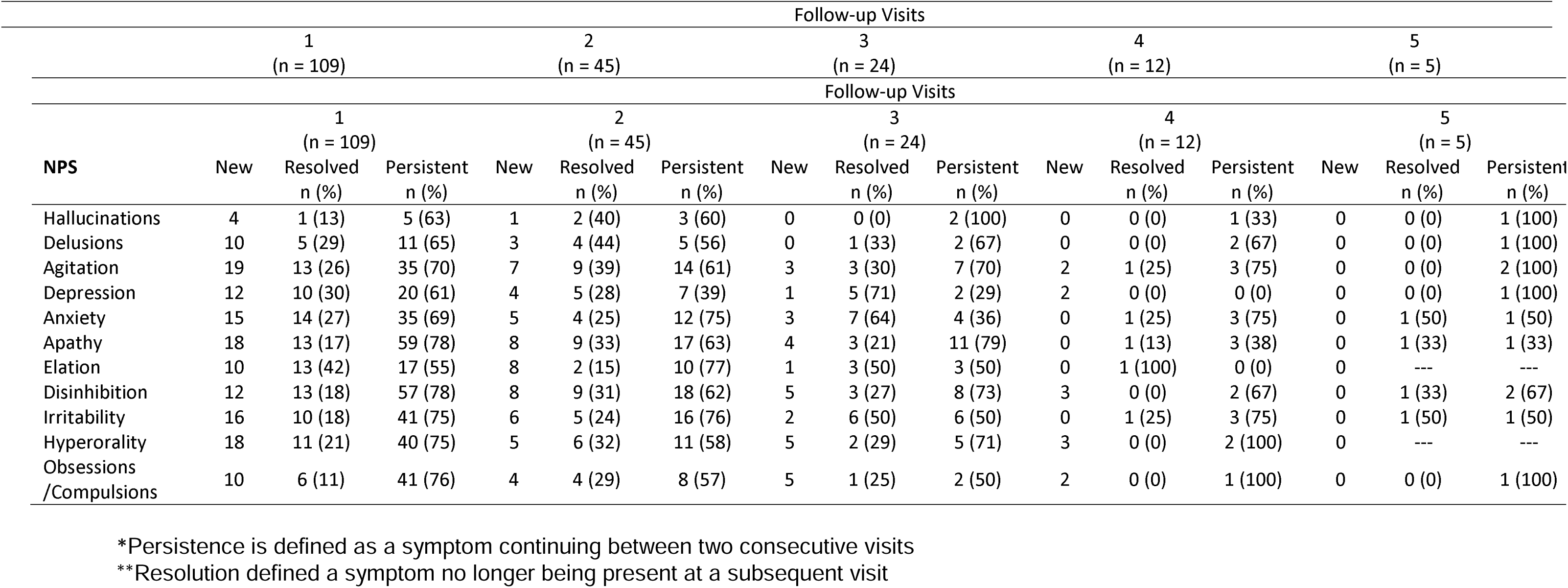
Persistence of Symptoms across study visits in bvFTD.

| Follow-up Visits |  |  |  |  |  |  |  |  |  |  |  |  |  |  |  |
| --- | --- | --- | --- | --- | --- | --- | --- | --- | --- | --- | --- | --- | --- | --- | --- |
| 1<br>(n = 109) |  |  | 2<br>(n = 45) |  |  | 3<br>(n = 24) |  |  | 4<br>(n = 12) |  |  | 5<br>(n = 5) |  |  |  |
| Follow-up Visits |  |  |  |  |  |  |  |  |  |  |  |  |  |  |  |
| NPS | 1<br>(n = 109) |  |  | 2<br>(n = 45) |  |  | 3<br>(n = 24) |  |  | 4<br>(n = 12) |  |  | 5<br>(n = 5) |  |  |
|  | New | Resolved<br>n (%) | Persistent<br>n (%) | New | Resolved<br>n (%) | Persistent<br>n (%) | New | Resolved<br>n (%) | Persistent<br>n (%) | New | Resolved<br>n (%) | Persistent<br>n (%) | New | Resolved<br>n (%) | Persistent<br>n (%) |
| Hallucinations | 4 | 1 (13) | 5 (63) | 1 | 2 (40) | 3 (60) | 0 | 0 (0) | 2 (100) | 0 | 0 (0) | 1 (33) | 0 | 0 (0) | 1 (100) |
| Delusions | 10 | 5 (29) | 11 (65) | 3 | 4 (44) | 5 (56) | 0 | 1 (33) | 2 (67) | 0 | 0 (0) | 2 (67) | 0 | 0 (0) | 1 (100) |
| Agitation | 19 | 13 (26) | 35 (70) | 7 | 9 (39) | 14 (61) | 3 | 3 (30) | 7 (70) | 2 | 1 (25) | 3 (75) | 0 | 0 (0) | 2 (100) |
| Depression | 12 | 10 (30) | 20 (61) | 4 | 5 (28) | 7 (39) | 1 | 5 (71) | 2 (29) | 2 | 0 (0) | 0 (0) | 0 | 0 (0) | 1 (100) |
| Anxiety | 15 | 14 (27) | 35 (69) | 5 | 4 (25) | 12 (75) | 3 | 7 (64) | 4 (36) | 0 | 1 (25) | 3 (75) | 0 | 1 (50) | 1 (50) |
| Apathy | 18 | 13 (17) | 59 (78) | 8 | 9 (33) | 17 (63) | 4 | 3 (21) | 11 (79) | 0 | 1 (13) | 3 (38) | 0 | 1 (33) | 1 (33) |
| Elation | 10 | 13 (42) | 17 (55) | 8 | 2 (15) | 10 (77) | 1 | 3 (50) | 3 (50) | 0 | 1 (100) | 0 (0) | 0 | --- | --- |
| Disinhibition | 12 | 13 (18) | 57 (78) | 8 | 9 (31) | 18 (62) | 5 | 3 (27) | 8 (73) | 3 | 0 (0) | 2 (67) | 0 | 1 (33) | 2 (67) |
| Irritability | 16 | 10 (18) | 41 (75) | 6 | 5 (24) | 16 (76) | 2 | 6 (50) | 6 (50) | 0 | 1 (25) | 3 (75) | 0 | 1 (50) | 1 (50) |
| Hyperorality | 18 | 11 (21) | 40 (75) | 5 | 6 (32) | 11 (58) | 5 | 2 (29) | 5 (71) | 3 | 0 (0) | 2 (100) | 0 | --- | --- |
| Obsessions<br>/Compulsions | 10 | 6 (11) | 41 (76) | 4 | 4 (29) | 8 (57) | 5 | 1 (25) | 2 (50) | 2 | 0 (0) | 1 (100) | 0 | 0 (0) | 1 (100) |

### Caregiver Burden and Functional Impact of NPS

The affective cluster and disinhibited cluster were associated with higher levels of caregiver burden (Table 8). Participants with agitation had a mean Zarit Burden Inventory (ZBI) score of 40.1 (95% CI 38.0 – 42.2) compared to those without agitation (35.0 [95% CI 33.0 – 37.1], p-value <0.001). Within the disinhibited cluster, those with elation (40.9 [95% CI 37.7 – 44.1] versus 36.7 [95% CI 35.0 – 38.3], p-value 0.02) and disinhibition (40.6 [95% CI 38.9 – 42.3] versus 31.5 [95% CI 28.9 – 34.1], p-value <0.001) had higher ZBI scores than those without elation or disinhibition.

**Table 8:** Impact of NPS on Caregiver Burden and Functional Abilities in bvFTD.

|  | ZBI |  |  | FAQ |  |  |
| --- | --- | --- | --- | --- | --- | --- |
|  | mean (SD) |  | p-value | Mean (SD) |  | p-value |
|  | - | + |  | - | + |  |
| Hallucinations | 37.3 (16.2) | 36.7 (17.4) | 0.85 | 18.2 (9.2) | 21.9 (7.6) | 0.04 |
| Delusions | 37.3 (16.2) | 39.3 (15.3) | 0.31 | 18.2 (9.2) | 20.6 (7.7) | 0.05 |
| Agitation | 35.0 (15.8) | 40.1 (16.0) | **<0.001** | 18.2 (9.9) | 19.0 (8.1) | 0.36 |
| Depression | 37.0 (16.6) | 38.6 (15.1) | 0.31 | 19.3 (9.2) | 17.0 (8.4) | 0.02 |
| Anxiety | 36.0 (15.8) | 39.9 (15.9) | 0.01 | 18.2 (9.6) | 19.1 (8.2) | 0.37 |
| Apathy | 33.5 (17.8) | 39.3 (15.1) | **<0.001** | 15.1 (10.3) | 19.6 (8.3) | **<0.001** |
| Elation | 36.7 (15.7) | 40.9 (16.8) | 0.02 | 18.7 (9.6) | 18.5 (7.4) | 0.85 |
| Disinhibition | 31.5 (16.1) | 40.6 (15.1) | **<0.001** | 17.8 (11.0) | 18.9 (8.0) | 0.26 |
| Irritability | 34.5 (16.1) | 40.3 (15.5) | **<0.001** | 18.4 (9.8) | 18.8 (8.3) | 0.73 |
| Hyperorality | 36.5 (15.6) | 39.5 (16.3) | 0.05 | 16.3 (8.7) | 21.2 (8.0) | **<0.001** |
| Obsessions /Compulsions | 36.5 (15.4) | 39.8 (16.2) | 0.03 | 16.7 (9.1) | 21.1 (7.7) | **<0.001** |

\*t-values for continuous variables, $\chi^2$ for binary variables

Apathy was also associated with higher levels of caregiver burden (39.3 [95% CI 37.6 – 40.9] versus 33.5 [95% CI 30.3 – 36.7], p-value <0.001).

Symptoms in the obsessive cluster (hyperorality, obsessions/compulsions) as well as apathy were associated with worse functional impairment (Table 8). Functional Activity Questionnaire (FAQ) scores for those with hyperorality were 21.2 [95% CI 20.2 – 22.3] versus 16.3 [95% CI 14.9 – 17.7] in those without hyperorality (p-value <0.001). Those with obsessions/compulsions had FAQ scores of 21.1 [95% CI 20.1 – 22.2] versus 16.7 [95% CI 15.2 – 18.1] in those without obsessions/compulsions (p-value <0.001). Those with apathy had FAQ scores of 19.6 [95% CI 18.6 – 20.5] compared to 15.1 [95% CI 12.9 – 17.4] in those without apathy (p-value <0.001).

## Discussion

### Overview

This study examined NPS in genetic and sporadic bvFTD, finding that NPS are common, cluster into distinct phenotypes, and have variable temporal stability. While NPS are common in many forms of neurodegenerative illness including AD, where prevalence of specific NPS often exceeds 40%, we show that in bvFTD multiple NPS are nearly universal with over 70% of participants experiencing three or more symptoms at baseline and over 90% experiencing at least one NPS at baseline.(30) As expected, distinct NPS clusters were identified in early and advanced stages of bvFTD: Affective (depression, anxiety, irritability, agitation); Disinhibited (elation, disinhibition, apathy); Compulsive (hyperorality, obsessive/compulsive behavior); Psychosis (hallucinations and delusions). As anticipated, NPS were shown to fluctuate over time, with elation, anxiety, and depression showing the most variability across visits.

### Neuropsychiatric Symptom Clusters

A primary goal of this study was to identify distinct clusters of NPS in bvFTD. The four NPS clusters that emerged in both early and advanced-stage disease overlap phenotypically with some features of primary psychiatric disorders (PPD), suggesting that shared neural network disruptions could underlie the emergence of specific symptoms. Future studies aimed at identifying functional and structural neural correlates of these NPS clusters could be useful in efforts to understand the neurobiological substrates, and to develop targeted therapies.

#### Affective Symptom Cluster

The affective symptom cluster identified in this study consisted of a combination of depression, anxiety, agitation, and irritability. Of these symptoms, anxiety, irritability, and agitation were among those most associated with elevated caregiver burden, suggesting that this cluster has a particularly significant impact on patient and caregiver quality of life. Current management of affective symptoms in bvFTD is challenging. Affective symptoms in bvFTD can be alleviated by treatment with selective serotonin reuptake inhibitors (SSRI), but responses are often incomplete—there are often residual symptoms despite treatment.(31) Alterations in 5-HT activity resulting from neuronal loss, tau deposition in the raphe nucleus and loss of 5-HT receptors in the midbrain, frontal, and temporal lobes have been associated with the development of affective symptoms in bvFTD.(32–34) TAR DNA-binding protein 43 (TDP-43) deposition in the CA3 subregion of the hippocampus has also been linked to depression in bvFTD. (34–36) Future therapies to target relevant networks impacted by these changes in bvFTD include non-invasive electrical brain stimulation and the use of small molecules acting as multi-modal serotonergic neuromodulators. Targeted therapy, directed at the underlying neurobiology, could be more effective than current treatments for affective symptoms in bvFTD, significantly improving patient and caregiver quality of life.

#### Disinhibited Symptom Cluster

The disinhibited symptom cluster identified in this study consisted of elation and disinhibition in early-stage participants and apathy and disinhibition in advanced-stage participants. The early-stage cluster has some symptom overlap with bipolar disorder, where dysfunction in the dorsal cognitive circuit and fronto-limbic circuit have been implicated in symptoms of hypomania and mania.(40) Interestingly, lithium—the gold standard for maintenance therapy in bipolar disorder—has been used with some success for the management of behavioral symptoms in bvFTD.(41) The observation that patients with bvFTD and those bipolar disorder can benefit from lithium treatment suggests similar neural network disruptions. Future studies are needed to clarify the specific neural networks impacted in this symptom cluster to inform design of targeted therapies and to determine which bvFTD patients exhibiting elation and disinhibition are the best candidates for therapies like lithium.

The advanced-stage bvFTD phenotype included disinhibition and apathy, which overlap with symptoms often seen in other neurodegenerative conditions. Impulse control disorder (ICD) in Parkinson’s disease (PD), for instance, can involve a variety of dysregulated behaviors including excessive gambling, sex, spending, and overeating, overlapping with symptoms seen in hypomania/mania. (42–44) Interestingly, however, patients experiencing ICD in PD also have elevated rates of apathy, with some hypothesizing that both ICD and apathy arise from dopaminergic dysfunction within the mesocorticolimbic pathway—a network that has been implicated in hypomania in bipolar disorder. (45, 46) Here, the similarity between symptoms observed in bvFTD and disorders with overlapping symptom profiles, highlights the possibility of shared neural network disruptions and the potential for targeted therapies.

The observation that MAPT mutation carriers were less likely to exhibit disinhibition compared to C9orf72 mutation carriers or sporadic bvFTD is also interesting and suggests that in addition to neural network dysfunctions, neuropathological states may influence NPS profiles in different ways. For instance, TDP-43 predominant neuropathology may be more likely to lead to disinhibition compared to tau-predominant pathologies. Further work is needed to correlate bvFTD-related NPS with both neural network dysfunction and neuropathological background.

#### Compulsive Symptom Cluster

The compulsive symptom cluster identified in this study consisted of hyperorality and ritualistic/obsessive-compulsive behavior. These symptoms were associated with high levels of functional disability, suggesting a high correlation with skills relevant to day-to-day functioning. Hyperorality and ritualistic/obsessive-compulsive behavior have been found to co-occur in previous studies of bvFTD and are associated with striatal gray matter volume loss. (47–49)These symptoms overlap with those seen in obsessive-compulsive disorder (OCD) and binge eating disorder (BED), both of which are associated with dysfunction in striatal circuits including the cortico-striatal-thalamic network. Obsessive symptoms are challenging to treat in both bvFTD and OCD, often with limited benefit from psychotropic medicines. However, targeted non-invasive brain stimulation techniques, stimulating the dorsolateral prefrontal cortex (dlPFC) as a means to modulate cortico-striato-thalamo-cortical circuits, have been used successfully in treating refractory symptoms of OCD and BED and may have significant potential in bvFTD. (50–52) The observation that MAPT mutation carriers were less likely to exhibit hyperorality or obsessions/compulsions compared to sporadic bvFTD is interesting and raises questions about how neuropathology influences NPS phenotypes.

While our sensitivity analysis including questionable cases of ritualistic/obsessive-compulsive behavior and hyperorality resulted in small changes to the structure of symptom clusters, given the goal of assessing symptoms that are clearly present as opposed to threshold or questionable cases, we felt the final analysis should include only those participants with symptoms marked as “definitively present.”

#### Psychosis Symptom Cluster

The psychosis symptom cluster in this study consisted of hallucinations and delusions. While relatively rare, these symptoms do occur in bvFTD, particularly in those with TDP-43 pathology.(53) Treatment of psychosis in bvFTD can be challenging as parkinsonism is not uncommon and amplifies the risk for drug-induced parkinsonism. Antipsychotics can be effective but, response rates are imperfect, and utility is limited by the potential for parkinsonism and other adverse effects. Psychotic symptoms in bvFTD have been correlated with grey matter atrophy in regions including the anterior insula, left thalamus, and cerebellum.(38) As previously described, extensive areas of reduced 5-HT activity are characteristic of bvFTD and could correspond with atrophy in areas associated with psychosis.

Pimavanserin, an antipsychotic with inverse agonist and antagonist properties at the 5-HT2A receptor, may have a future role in treating psychosis in bvFTD by modulating relevant circuit dysfunction, representing another example where identifying neural correlates of NPS clusters may facilitate the development of more effective, targeted therapies for NPS in bvFTD.(54)

### Neuropsychiatric Symptom Fluctuations

A complementary aim of this study was to evaluate the stability of specific NPS over time. A major priority in advancing care for patients with bvFTD is finding ways to optimize clinical trial participation for emerging disease-modifying therapies. Identifying patients at the earliest sign of symptoms, prior to significant neurodegeneration, could maximize the potential to favorably alter the disease course. Most trials rely on measures like the CDR® plus NACC FTLD to assess disease severity for the purpose of trial enrollment. Scales like the CDR® plus NACC FTLD have many strengths but do not fully describe the heterogeneity of the earliest stages of bvFTD. In other words, psychiatric phenomena may be relevant to preclinical bvFTD constructs.

Whether psychiatric symptoms outside the core diagnostic criteria should be included in bvFTD severity rating scales is an important and open question. Ideal elements for a disease severity rating scale are symptoms that reflect neurodegeneration and are sensitive to the temporal progression. In this study, we observed that some NPS, particularly those that are already included in the core bvFTD diagnostic criteria (disinhibition, apathy, compulsions, hyperorality) are relatively stable, persisting across visits. However, other NPS outside the core diagnostic criteria—including depression, anxiety, and elation— are less stable. Psychotic symptoms appear to be relatively stable. Given these findings, some psychiatric symptoms outside the core behavioral criteria may not reflect neurodegeneration reliably and therefore, would not be considered indexes of disease severity. Further longitudinal work is needed to definitively establish the role of psychiatric symptoms in the staging and monitoring of bvFTD progression.

While our study benefits from a large well-characterized population with expert diagnoses of bvFTD, several limitations merit consideration. Our study relies on clinical as opposed to pathological diagnosis, and so the possibility of diagnostic error, i.e., that a proportion have other neurodegenerative diseases (AD, DLB, PPD, etc.) cannot be discounted. This problem is mitigated by the study’s reliance on expert clinicians establishing diagnosis using standard criteria in a formal consensus conference process. Another caveat is that the analyses do not take into account NPS severity; relying on a binary assessment of NPS (presence/absence) does not fully characterize psychiatric symptoms or their severity. In addition, assessments for many NPS domains were based on informant report which may not be as reliable as a formal clinical interview. Future studies will benefit from the use of more sophisticated assessments, including structured psychiatric interviews, to provide deeper phenotyping of bvFTD-related psychiatric phenomena. In addition, our use of factor analysis to define NPS clusters does not allow for a definitive examination of the neurobiology underlying psychiatric phenotypes in bvFTD. While useful for hypothesis generation, our findings will be strengthened by future studies that prospectively examine the functional and structural neural correlates of NPS in bvFTD. Our assessment of symptom fluctuations also did not account for any pharmacologic or non-pharmacologic interventions. Future studies will benefit from a comprehensive record of treatments prescribed, allowing for better characterization of the natural history of bvFTD-related NPS and the efficacy of existing interventions. Finally, our study population is limited to participants recruited for participation at academic centers and consisted predominantly of white participants living in North America. This limits the generalizability of our results to other ethnocultural groups and geographic locations.

## Conclusion

In this study, we show that NPS in bvFTD cluster into four distinct domains, with symptoms overlapping those seen in a variety of primary psychiatric conditions and other neurodegenerative disorders. We highlight the need for future work utilizing prospective samples with well characterized psychiatric phenomena to identify the brain (structural and functional) correlates. We also observed that NPS have temporal variability, suggesting that while neuropsychiatric symptoms are central to the clinical presentation and lived experience of bvFTD, they may not be reliable markers of disease progression. Overall, our study highlights the potential for advancing treatment of psychiatric symptoms in bvFTD. Future studies are needed to clarify the early functional neuroanatomic changes accompanying these psychiatric phenotypes, as well as the neuropathological substrates, in order to inform the development of novel pharmacotherapies and methods for targeted neuromodulation.

## Author Contributions

All authors contributed to the design and/or analytic approach of the study. CM performed initial analyses and drafted the manuscript text. All other authors reviewed each draft and made substantive revisions and approved the final manuscript.

## Conflicts of Interest

Preliminary analysis from this work was presented as an abstract at the ISFTD conference in Amsterdam, NL on September 21, 2024. CM was funded by KL2TR003099 and K23AG088248. AP is funded by NIA/NINDS (K23AG059891 and U01NS102035). JR is funded by NIA/NIH K23AG059888, AlzOut, Shenandoah fund and Jon and Gale Love Alzheimer’s fund and is a site PI for clinical trials sponsored by Eli-Lilly and Eisai and a consultant and speaker for Roon Health, Inc. ZKW is partially supported by the NIH/NIA and NIH/NINDS (1U19AG063911, FAIN: U19AG063911), Mayo Clinic Center for Regenerative Medicine, the gifts from the Donald G. and Jodi P. Heeringa Family, the Haworth Family Professorship in Neurodegenerative Diseases fund, The Albertson Parkinson’s Research Foundation, and PPND Family Foundation. He serves as PI or Co-PI on Biohaven Pharmaceuticals, Inc. (BHV4157-206) and Vigil Neuroscience, Inc. (VGL101-01.002, VGL101-01.201, PET tracer development protocol, Csf1r biomarker and repository project, and ultra-high field MRI in the diagnosis and management of CSF1R-related adult-onset leukoencephalopathy with axonal spheroids and pigmented glia) projects/grants. He serves as Co-PI of the Mayo Clinic APDA Center for Advanced Research and as an external advisory board member for the Vigil Neuroscience, Inc., and as a consultant on neurodegenerative medical research for Eli Lilli & Company. IL’s research is supported by the National Institutes of Health grants: 2R01AG038791-06A, U01NS100610, R25NS098999; U19 AG063911-1 and 1R21NS114764-01A1; the Michael J Fox Foundation, Parkinson Foundation, Lewy Body Association, CurePSP, Roche, Abbvie, Biogen, Lundbeck, EIP-Pharma, Biohaven Pharmaceuticals, Novartis, and United Biopharma SRL, UCB. She is a member of the Scientific Advisory Board for Amydis but does not receive funds and from the Rossy PSP Program at the University of Toronto. She receives her salary from the University of California San Diego and as Chief Editor of Frontiers in Neurology. VK is partially supported by the NIH/NIA and NIH/NINDS (R01AG064093, R01NS108452). AMS has received research support from the NIA/NIH, Bluefield Project to Cure FTD, the Alzheimer’s Association, the Larry L. Hillblom Foundation, and the Rainwater Charitable Foundation, and has provided consultation to Alector, Eli Lilly/Prevail Therapeutics, Passage Bio, and Takeda. C.U.O. has received research funding from the NIH, Lawton Health Research Institute, National Ataxia Foundation, Alector and Transposon. He is also supported by the Robert and Nancy Hall Brain Research Fund, the Jane Tanger Black Fund for Young-Onset Dementias and a gift from Joseph Trovato. He is a consultant with Alector Inc., Acadia Pharmaceuticals, Reata Pharmaceuticals, Otsuka Pharmaceuticals, Lykos Therapeutics, and Zevra Therapeutics. He serves of the scientific advisory boards of the Tau Consortium and the FTD Disorders Registry. KT is supported by NIH/NIA (R01AG068881, R01AG075111 and R01AG075404). N.G. has participated or is currently participating in clinical trials of anti-dementia drugs sponsored by Bristol Myers Squibb, Eli Lilly/Avid Radiopharmaceuticals, Janssen Immunotherapy, Novartis, Pfizer, Wyeth, SNIFF (The Study of Nasal Insulin to Fight Forgetfulness) and the A4 (The Anti-Amyloid Treatment in Asymptomatic Alzheimer’s Disease) trial. She receives research support from Tau Consortium and the Association for Frontotemporal Dementia and is funded by the NIH.

**Supplementary Figure 1:**
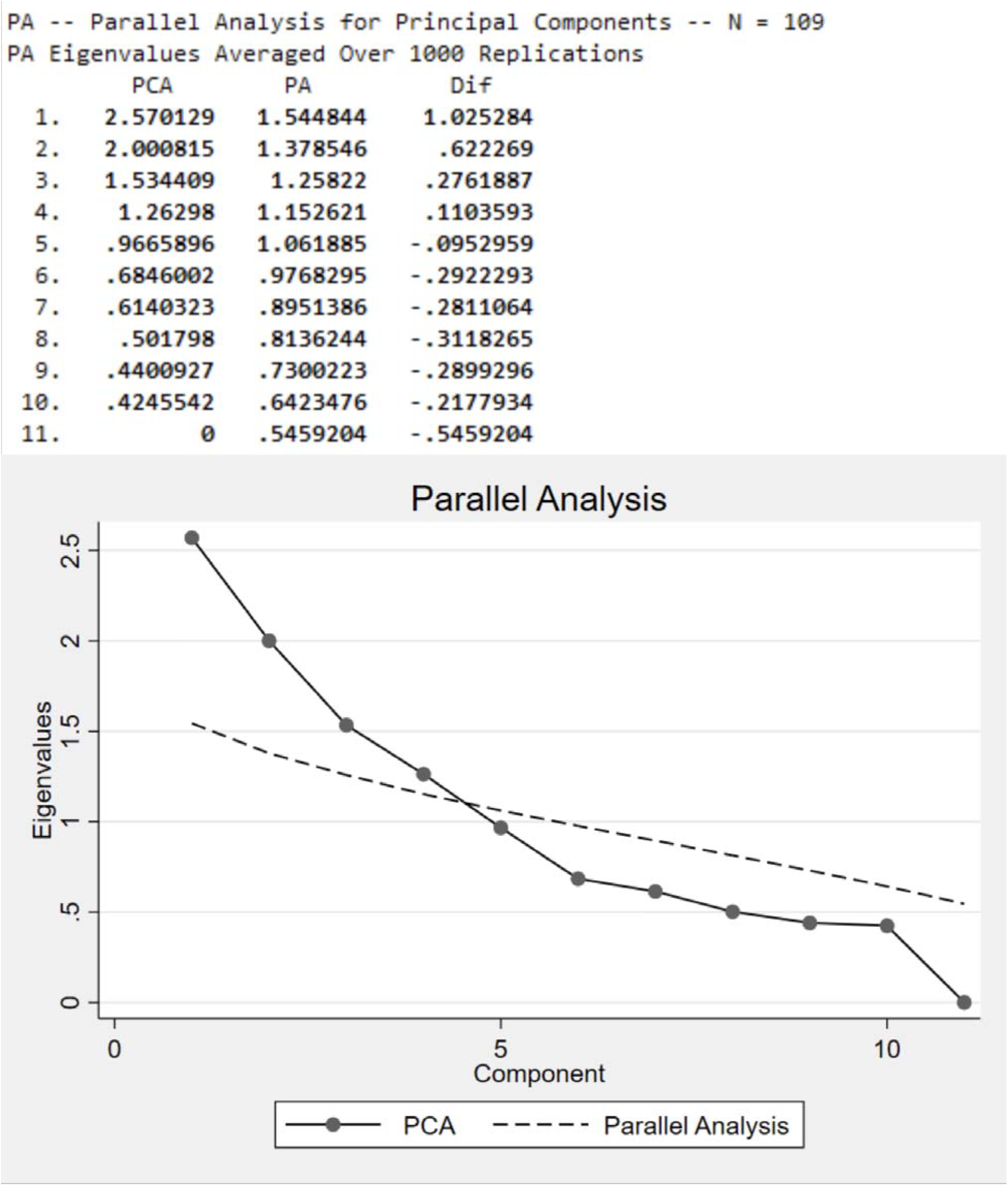
Four-Factor NPS Cluster Model in Early-Stage FTD.

**Supplementary Figure 2:**
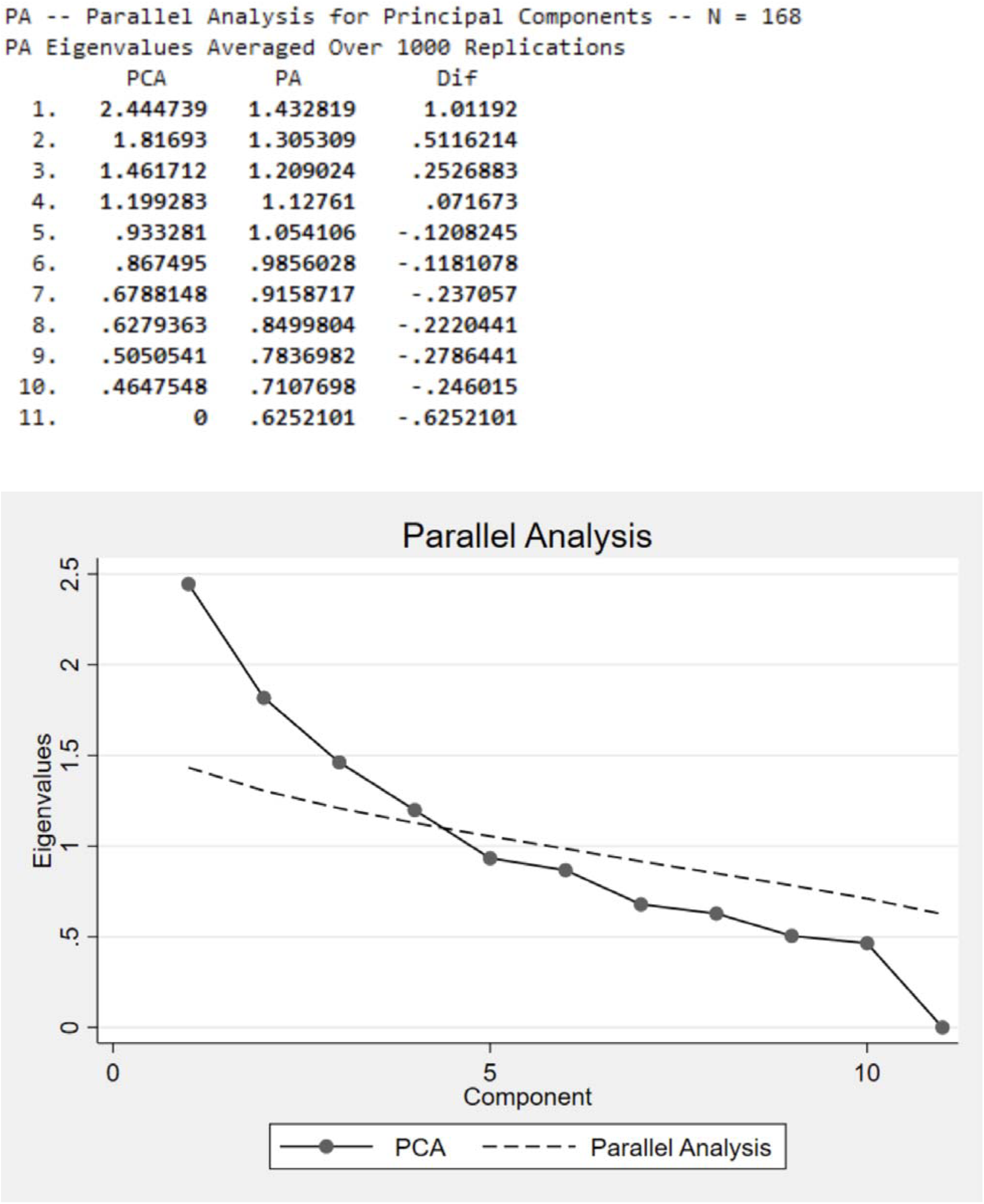
Four-Factor NPS Cluster Model in Advanced-Stage FTD.

**Supplementary Figure 3:**
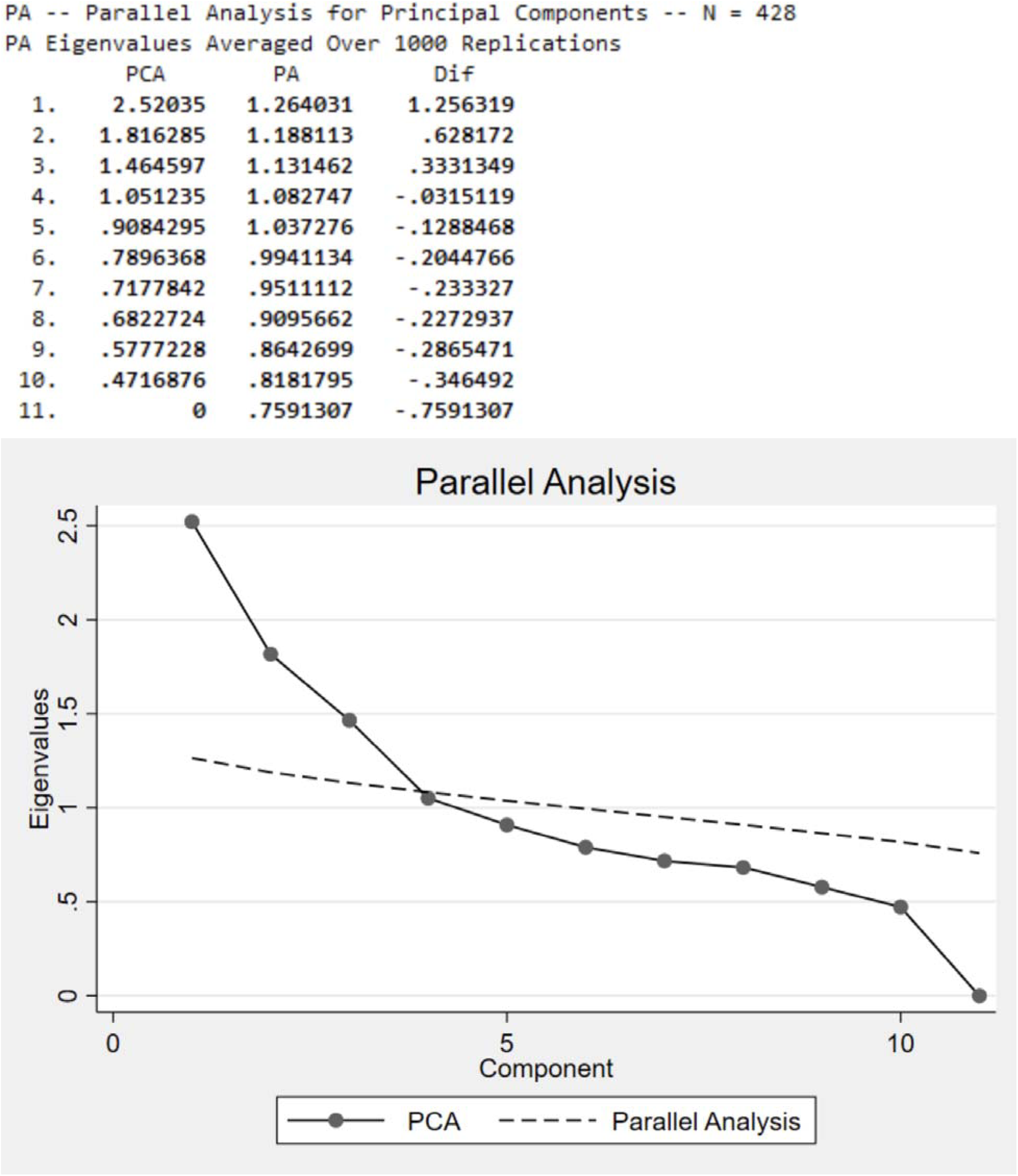
Four-Factor NPS Cluster Model Across All Visits and Disease Severity.

## Data Availability

All data produced are available online at https://www.allftd.org

